# A Universal Day Zero Infectious Disease Testing Strategy Leveraging CRISPR-based Sample Depletion and Metagenomic Sequencing

**DOI:** 10.1101/2022.05.12.22274799

**Authors:** Agnes P. Chan, Azeem Siddique, Yvain Desplat, Yongwook Choi, Sridhar Ranganathan, Kumari Sonal Choudhary, Josh Diaz, Jon Bezney, Dante DeAscanis, Zenas George, Shukmei Wong, William Selleck, Jolene Bowers, Victoria Zismann, Lauren Reining, Sarah Highlander, Yaron Hakak, Keith Brown, Jon R. Armstrong, Nicholas J. Schork

## Abstract

The lack of preparedness for detecting the highly infectious SARS-CoV-2 pathogen, the pathogen responsible for the COVID-19 disease, has caused enormous harm to public health and the economy. It took ∼60 days for the first reverse transcription quantitative polymerase chain reaction (RT-qPCR) tests for SARS-CoV-2 infection developed by the United States Centers for Disease Control (CDC) to be made publicly available. It then took >270 days to deploy 800,000 of these tests at a time when the estimated actual testing needs required over 6 million tests per day. Testing was therefore limited to individuals with symptoms or in close contact with confirmed positive cases. Testing strategies deployed on a population scale at ‘Day Zero’ i.e., at the time of the first reported case, would be of significant value. Next Generation Sequencing (NGS) has such Day Zero capabilities with the potential for broad and large-scale testing. However, it has limited detection sensitivity for low copy numbers of pathogens which may be present. Here we demonstrate that by using CRISPR-Cas9 to remove abundant sequences that do not contribute to pathogen detection, NGS detection sensitivity of COVID-19 is comparable to RT-qPCR. In addition, we show that this assay can be used for variant strain typing, co-infection detection, and individual human host response assessment, all in a single workflow using existing open-source analysis pipelines. This NGS workflow is pathogen agnostic, and therefore has the potential to transform how both large-scale pandemic response and focused clinical infectious disease testing are pursued in the future.

**SIGNIFICANCE STATEMENT:** The lack of preparedness for detecting infectious pathogens has had a devastating effect on the global economy and society. Thus, a ‘Day Zero’ testing strategy, that can be deployed at the first reported case and expanded to population scale, is required. Next generation sequencing enables Day Zero capabilities but is inadequate for detecting low levels of pathogen due to abundant sequences of little biological interest. By applying the CRISPR-Cas system to remove these sequences *in vitro*, we show sensitivity of pathogen detection equivalent to RT-qPCR. The workflow is pathogen agnostic, and enables detection of strain types, co-infections and human host response with a single workflow and open-source analysis tools. These results highlight the potential to transform future large-scale pandemic response.

## INTRODUCTION

The current COVID-19 pandemic has exposed the threat of infectious diseases to human health and safety and the essential role that pandemic preparedness can play in combatting such threats(1). The deaths directly attributed to COVID-19 exceed 6.1M globally with over 927,000 deaths in the United States (US), making COVID-19 the third most lethal viral pandemic in the past century, behind the Spanish Flu of 1918 and HIV (2). National and international pandemic preparedness plans are essential, as COVID-19 is likely not the last pandemic in our future. As an indication of why pandemic preparedness is so important, consider that the first confirmed case of SARS-CoV2, the cause of the COVID-19 pandemic, occurred around December 1, 2019 (3). From that time, termed ‘Day Zero,’ it took ∼42 days before the SARS-CoV-2 viral genome sequence was publicly released (4). One month later, the CDC produced 90 initial testing kits, which unfortunately had documented contamination issues (5). Over the course of the next 6-8 months, the US was able to produce approximately 800,000 tests per day, when the estimated need at the time was an ability to test six million individuals per day (6). Because of limited capacity, testing was recommended only to those individuals with symptoms or in close contact with confirmed positive COVID-19 cases, leaving asymptomatic carriers as a major source of transmission. By July 2020, over 350,000 US residents had confirmed infections with over 4,500 confirmed deaths (2).

With these facts in mind, the US government has proposed a 10-year set of activities and $65B in funding for pandemic preparedness balanced with a belief of a strong return on investment given the ∼$16-Trillion economic impact in the US of the COVID-19 pandemic over the past 24 months (1). With a growing global population, increased access to global travel, encroachment on previously less populated locations, the increasing number of labs researching infectious disease, and the potential for nefarious intent to weaponize biological pathogens, this modest investment seems more than justified.

What is necessary to enable the proposed vision of pandemic preparedness are testing and response strategies that are deployable at Day Zero to combat any future pathogen outbreak before it progresses to a pandemic. Such an approach, by necessity, needs to be pathogen agnostic and, ideally, would provide more detailed information about an individual’s reaction than the mere presence of a pathogen. NGS can fulfill those requirements. Shotgun NGS generates sequence information from every molecule in a sample (e.g., nasal swab, saliva, blood), and has shown to be applicable for human infectious disease diagnostic tests (7–9), respiratory infections (10), and universal pathogen detection (11–15). Prior to the pandemic, an NGS-based study involving a population of ∼1,000 pangolins from a wet market in Wuhan China, identified coronavirus infections in over 70% of the pangolin samples tested (16). The authors sequenced total RNA from pangolin samples and achieved similar genome characterizations using *de novo* assembly and reference-based methods, ultimately showing that new viruses can be identified by such an approach. Advances in sequencing technology, reductions in sequencing cost and increases in instrument throughput, the advent of portable instruments, and internet connectivity and cloud-based data analysis means that NGS technology has reached a point where it is capable of the speed and advanced data processing necessary for Day Zero deployment and of handling population-scale throughput in response to a pandemic.

However, there are caveats to the use and deployment of NGS as part of a pandemic preparedness strategy. First, NGS has not been applied for routine clinical use for infectious disease diagnosis. This is because many sequencers have been housed for the most part in academic research institutions and private research organizations that lack the logistical infrastructure necessary for sample attainment, upstream processing, return of results and biosecurity level clearance to handle samples with active virus. However, with the acceptance of ‘at home’ sample collection and telehealth consultation via the internet, spurred by the need to keep individuals distanced from others, particularly from overburdened hospitals, the logistical infrastructure is far better than it has been before. Second, there has been a reluctance to implement NGS in hospital settings because of the amount of data generated and the computational infrastructure required to run sequence data processing pipelines. However, complex computational analysis is only needed until the pathogen of interest is identified and a reference genome of that pathogen is assembled, since mapping reads to a reference for identification purposes is relatively straightforward. Third, traditional NGS-based metagenomic tests for COVID-19 are inefficient because of relatively large sample input requirements and lower sensitivity for the virus due to overabundant, uninformative nucleic acid molecules from the human host and/or common commensal microbes.

Here we present a molecular enrichment strategy to overcome these limitations in the use of NGS protocols, by using a Clustered Regularly Interspersed Short Palindromic Repeats (CRISPR-Cas9) system to specifically target and remove abundant host and common human microbial sequences (17–19). This CRISPR-NGS technology has been patented and commercialize under the CRISPRclean^®^ name by Jumpcode Genomics, Inc. The technology uses a multiplexed pool of single guide RNAs (sgRNA) in a ribo-nucleoprotein complex (RNP) formed with the Cas9 enzyme. Double stranded cuts are generated in undesired NGS molecules, which is then followed by adapter specific PCR to enrich for un-cut NGS library molecules for enhanced sequencing. To evaluate the performance of the technology, we performed two critical assessment steps of COVID-19 clinical specimens (i.e., nasal swabs): (1) we defined the human and bacterial ribosomal RNA (rRNA) compositions to set baselines for the assessment of rRNA depletion efficiency, and (2) we established the pathogen-microbiome compositions, using orthogonal bioinformatics protocols to assess taxonomic classification confidence. We show that SARS-CoV-2 detection sensitivity using this method is comparable to RT-qPCR based detection for samples with C_t_ values up to 35. We also demonstrate that the CRISPR-NGS strategy enables variant strain typing, detection of co-infecting agents, identification of antimicrobial resistance genes (AMR), and the reporting of human host responses to infection. Furthermore, using contrived samples containing viral nucleic acid, we show that the CRISPR-NGS approach can successfully detect other pathogens (e.g., Zika virus). A long-term vision is that the application of CRISPR-NGS technology accelerates patient diagnosis and preventive strategies of existing and future pathogens.

## RESULTS

### Study design and samples used for sequencing studies

Two types of samples were analyzed in this study: clinical specimens and contrived samples. For clinical specimens, human nasal swabs with COVID-19 infection status determined by RT-qPCR were previously collected from two locations, one in California and one in Arizona (referred to as site A and site B, respectively), and then processed and sequenced at separate sites (Jumpcode Genomics, San Diego, CA for site A samples and TGen, Phoenix, AZ for site B samples). In total, 56 patient specimens with confirmed positive COVID-19 status and 16 specimens with no detectable SARS-CoV-2 were analyzed in this study.

The contrived samples were generated by combining known viral RNA pathogens with human RNA. A viral reference genome mix, consisting of four RNA viruses (Zika virus, mammalian orthoreovirus, influenza B virus, human orthopneumovirus (i.e., respiratory syncytial virus, RSV) and SARS-CoV-2) was spiked at various concentrations into human lung total RNA samples. The samples contained a 10-fold dilution series of the viral reference mix, with titers ranging from an estimated 20 copies of each viral RNA genome to 20,000 copies. Various negative controls were also prepared, including human lung total RNA only (a no-viral RNA control) and a water-only sample (no-template control). The latter was included to monitor background contaminants that may originate from molecular reagents and NGS workflows. An overview of the workflow is shown in Figure 1.

**Figure 1.**
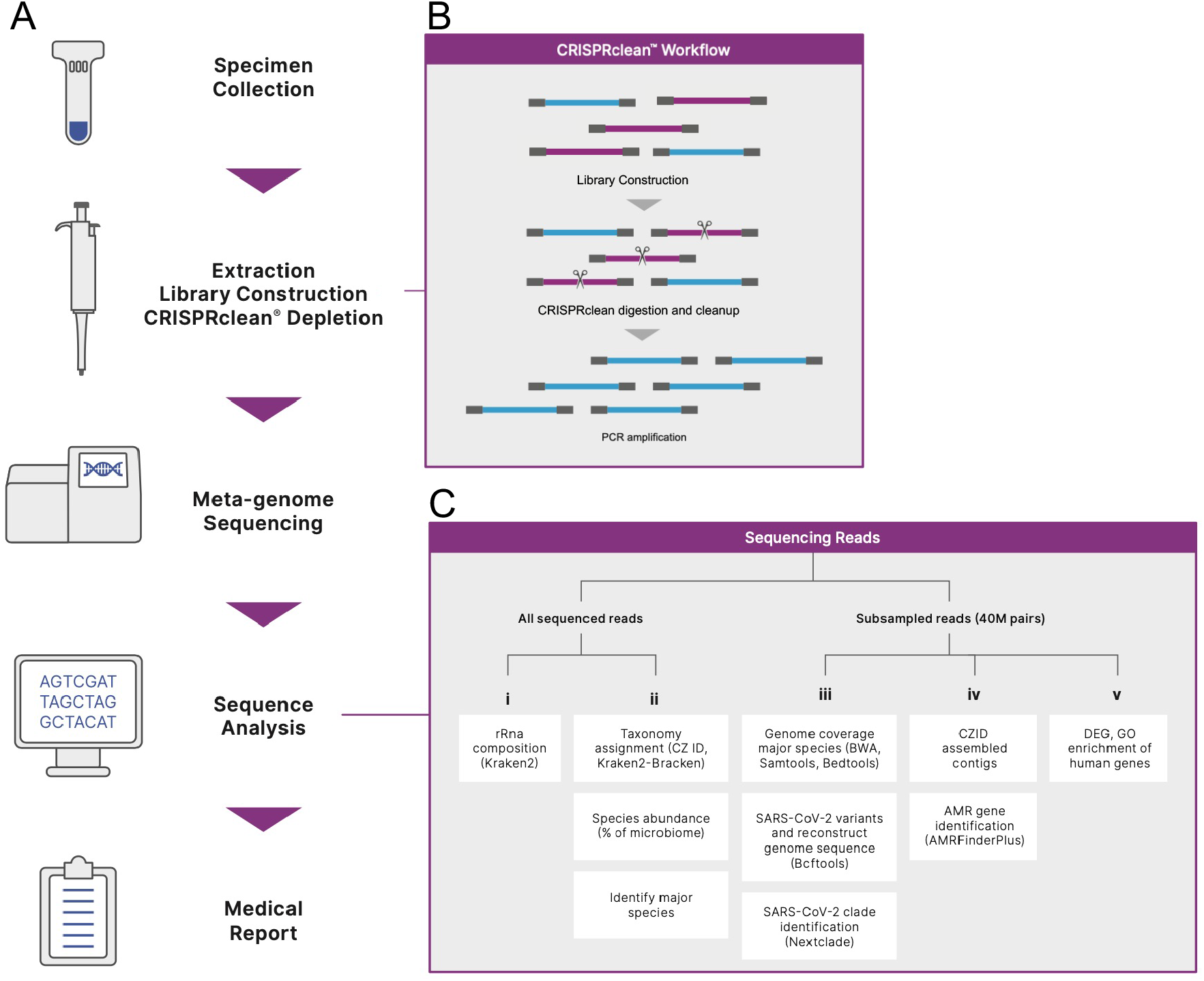
Study workflow and design. (A) From clinical specimens to reporting. (B) CRISPclean workflow. (C) Data analysis workflow: i. Estimating rRNA composition, ii. Reporting taxonomic classification and the abundance of pathogens and coinfections as percentage of microbiome (non-human) reads, iii. Reporting pathogen genome coverage metrics, iv. Reporting AMR, and v. Reporting host gene expression.

Both mock depletions (depletion without Cas9 and guide RNA) and Cas9 depletions were performed with site A specimens. Mock-depletion was not performed with site B specimens. A summary of sequencing statistics is provided in Supporting Table 1.

**Table 1.**
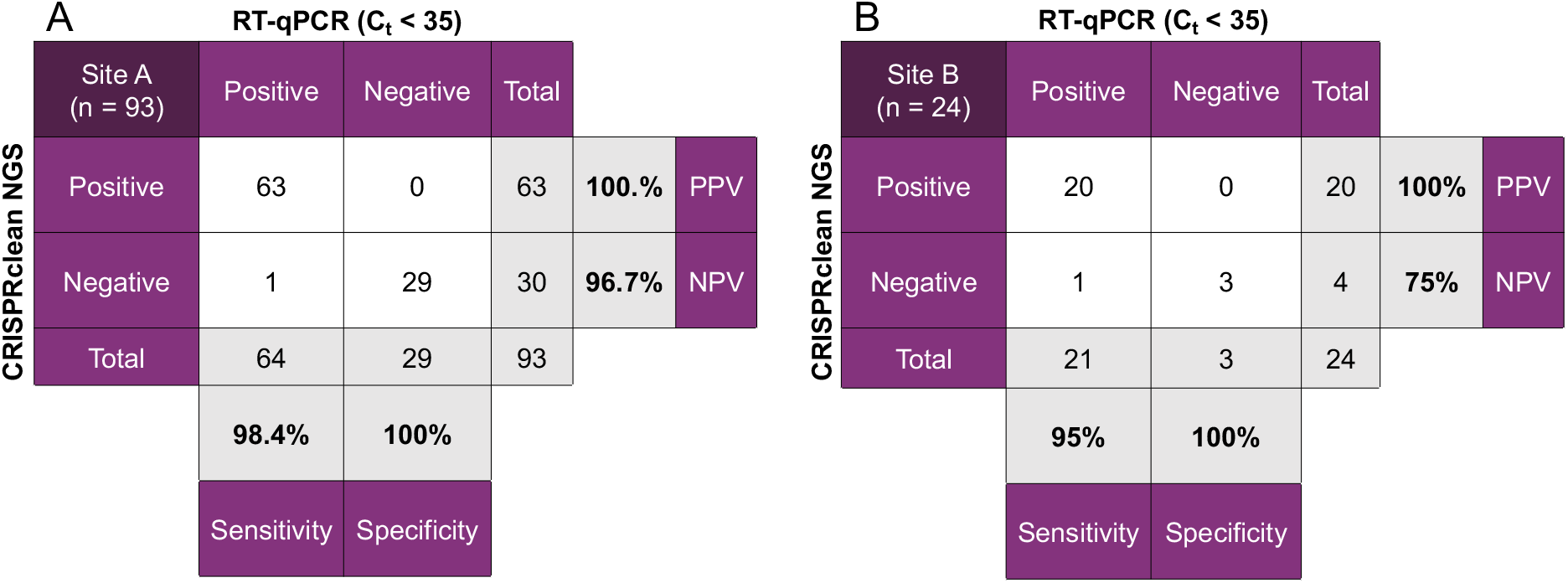
Contingency tables comparing the performance of CRISPRclean NGS and RT-qPCR for Site A and Site B. Samples with Ct < 35 were processed with the CRISPRclean NGS assay and positive/negative results compared to RT-qPCR results from the same samples. Sensitivity, specificity, positive predictive value (PPV) and negative predictive value (NPV) were calculated for both Site A and B. CRISPRclean results are comparable those seen from RT-qPCR.

| A | | RT-qPCR ($C_t < 35$) | | | | | |
| --- | --- | --- | --- | --- | --- | --- | --- |
| CRISPRclean NGS | Site A<br>(n = 93) | Positive | Negative | Total |  |  |  |
|  | Positive | 63 | 0 | 63 | 100.0% | PPV |  |
|  | Negative | 1 | 29 | 30 | 96.7% | NPV |  |
|  | Total | 64 | 29 | 93 |  |  |  |
|  |  | 98.4% | 100% |  |  |  |  |
|  |  | Sensitivity | Specificity |  |  |  |  |

Table 1. Contingency tables comparing the performance of CRISPRclean NGS and RT-qPCR for Site A and Site B.
| B | | RT-qPCR ($C_t < 35$) | | | | | |
| --- | --- | --- | --- | --- | --- | --- | --- |
| CRISPRclean NGS | Site B<br>(n = 24) | Positive | Negative | Total |  |  |  |
|  | Positive | 20 | 0 | 20 | 100% | PPV |  |
|  | Negative | 1 | 3 | 4 | 75% | NPV |  |
|  | Total | 21 | 3 | 24 |  |  |  |
|  |  | 95% | 100% |  |  |  |  |
|  |  | Sensitivity | Specificity |  |  |  |  |

### Ribosomal RNA landscape in clinical specimens

A key aspect of our NGS-based strategy is the use of CRISPR-Cas9 to remove ribosomal RNA sequences. To determine the effectiveness of rRNA removal, we set out to first establish the rRNA composition of the clinical nasal swab specimens by classifying sequence reads as bacterial, viral, or eukaryotic, and rRNA or non-rRNA (presumably mRNA), using Kraken2. For site A clinical specimens, the average content of human and bacterial rRNA in non-depleted NGS libraries was 63% and 0.85%, respectively, of total reads. After CRISPR-based rRNA depletion, human rRNA was detected at 0.74% and bacterial rRNA at 0.02% of total reads (Figure 2, Supporting Table 2). This indicates that 98% of bacterial rRNA and 99% of human rRNA were successfully depleted as a result of CRISPR-based depletion. Because of depletion, human non-rRNA, bacterial non-rRNA, and viral sequences were enriched by an average of 3.6-fold, 3-fold, and 6-fold, respectively, across all CRISPR-depleted samples. Each of these enriched sequence categories is important because they contribute to the downstream genomic characterization of SARS-CoV-2, co-infection pathogens, and host gene expression as discussed below. However, non-ribosomal human RNA molecules that have little to do with the patient response to infection may be candidates for further depletion.

**Figure 2.**
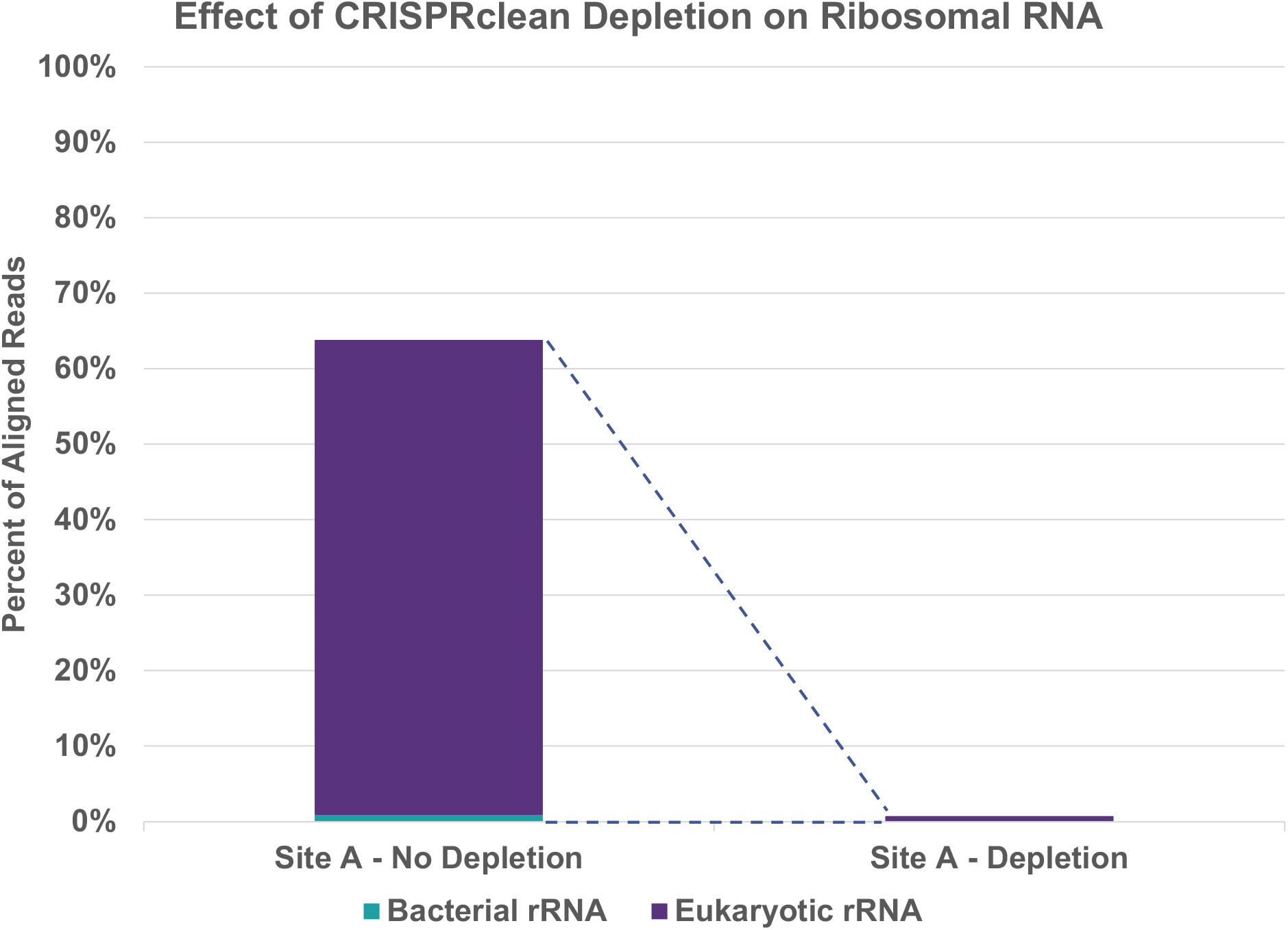
Ribosomal RNA composition before and after CRISPRClean depletion at Site A. The average percent of ribosomal aligned reads (y-axis) was determined for bacterial (blue) and eukaryotic ribosomal (purple) in all sample libraries from site A (n=180). Percent of aligned reads is shown with and without CRISPRclean depletion. CRISPRclean depletion removes nearly all bacterial and eukaryotic RNA.

Interestingly, the post-depletion human and bacterial rRNA profiles of site B samples were distinct from those of site A. The rRNA content of the non-depleted libraries from site B could not be determined because mock-depletions were not performed at site B. However, the library composition of human rRNA, bacterial rRNA, and virus after depletion was 4.8%, 16%, and 11%, respectively (Supporting Table 3). The reason for the higher proportion of bacterial rRNA after depletion in site B libraries relative to site A libraries is unclear but could be related to sample type or different methods of specimen collection, processing, and RNA extraction. Further investigation is warranted, for example, to determine whether the non-depleted bacterial rRNA sequences fall within or outside of the CRISPR guide RNA set of sequences used for rRNA depletion. In summary, the CRISPR-based rRNA depletion method presented here can achieve near-complete removal of human and bacterial rRNAs from clinical nasal swab specimens.

### Pathogen-microbiome landscape in clinical specimens and contrived samples

Next, we determined the pathogen-microbiome composition of the COVID-19 positive clinical specimens by focusing on the microbiome sequence read space of each specimen. Two independent and orthogonal bioinformatic approaches, the alignment/assembly-based Chan Zuckerberg ID (CZID) workflow (formerly known as IDSeq) (20) and the kmer-based Kraken2-Bracken workflow (21–23), were used for taxonomic classifications. For this analysis, all sequence reads generated for each specimen were analyzed without subsampling. The concordance of taxonomic assignments by the two different bioinformatics methods were also determined to identify taxa with confidence.

Analysis of the nasal swab clinical specimens using CZID showed that SARS-CoV-2 sequences were detected on average at 88,831 RPM and 192,582 RPM from site A and B, respectively (i.e., 8.8% and 19% of the nasal microbiome; Supporting Table 4). The highest SARS-CoV-2 viral load was detected at 983,494 RPM (98% of the nasal microbiome). Several respiratory pathogens were detected in site A specimens, including rhinovirus and Gammapapillomavirus. Rhinovirus A (232,730 RPM; 23%), rhinovirus B (2,920 RPM; 0.3%), rhinovirus C (40,840 RPM, 4%) were detected in individual specimens, and Gammapapillomavirus 1 in 16 specimens (e.g., 4,081 RPM; 0.4%). Respiratory tract commensals in descending order of abundance, as reported by the CZID workflow, from site A specimens include *Cutibacterium acnes* (226,086 RPM; 23%), *Corynebacterium segmentosum* (83,149 RPM; 8.3%), *Dolosigranulum pigrum* (29,396 RPM; 2.9%), *Staphylococcus epidermidis* (5562 RPM; 0.6%), and *Staphylococcus aureus* (1455 RPM; 0.15%). Among site B specimens, *Prevotella melaninogenica* (66,080 RPM; 6.6%), *Rothia mucilaginosa* (30,449 RPM; 3%), and *Prevotella jejuni* (19,838 RPM; 1.9%) were reported (Figure 3, Supporting Table 4). A commensal bacterium shared between site A and B specimens and detected at a comparable proportion/abundance is *Moraxella catarrhalis* (site A: 6,529 RPM, 0.7%; site B: 5,125 RPM, 0.5%).

**Figure 3.**
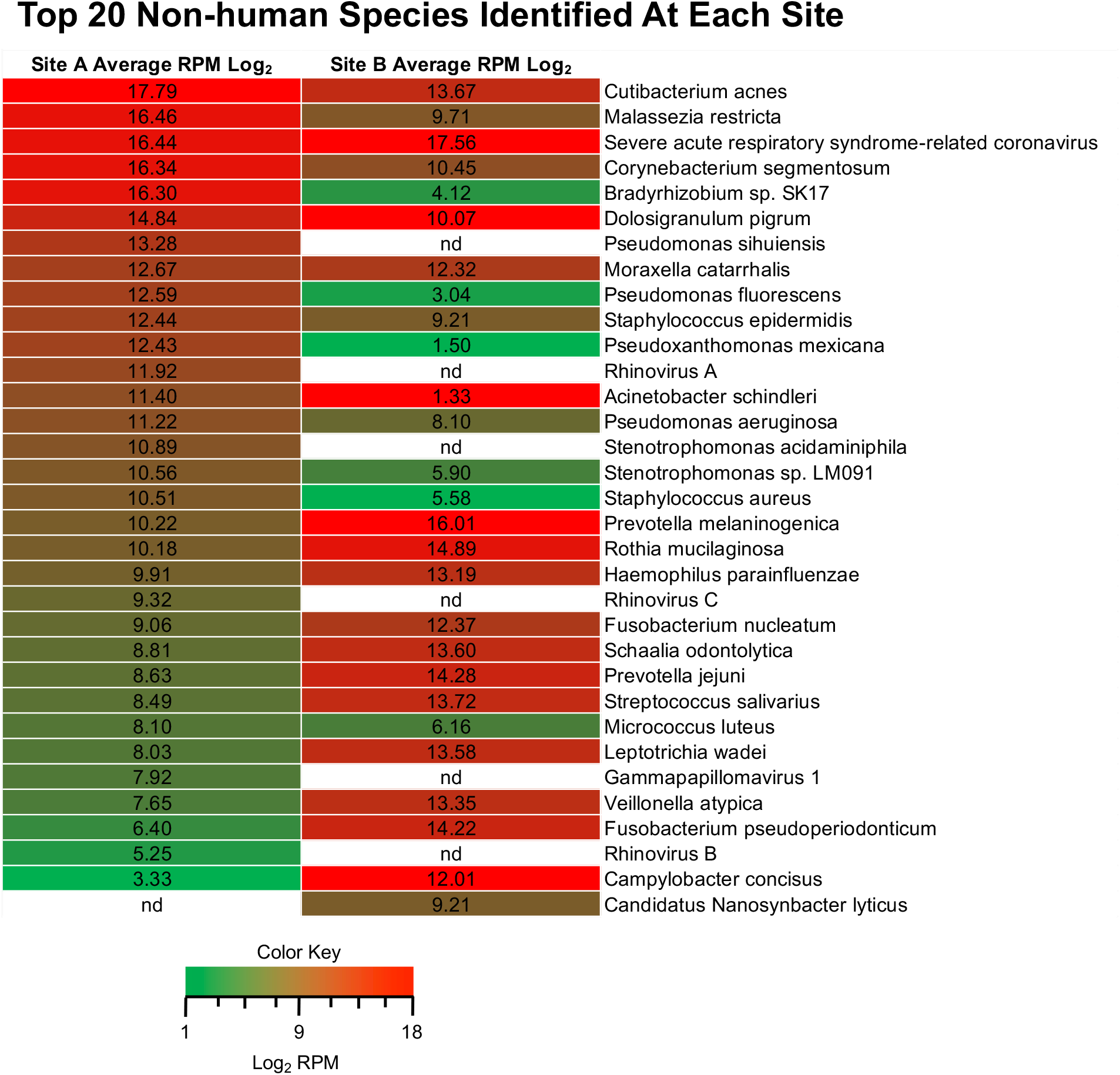
Heatmap of top 20 non-human species identified at Site A and B. The average log_2_ reads per million sequences (RPM) were calculated for non-human species in clinical samples from both sites and the top 20 species were identified from Site A (left column) and Site B (right column). Log_2_ RPM values are shown in each cell. Red represents higher and green represent lower log_2_ RPM values. Species not identified are in white (nd).

A comparison of the RPM values reported by the two classification methods, CZID and Kraken2-Bracken, showed concordance (within an absolute log_2_FC value of 2) for several bacterial residents of the respiratory tract and provided confidence regarding the taxonomic assignments (13 concordant – site A; 12 concordant – site B). For example, the log_2_FC of the CZID and Kraken2-Bracken RPM values of *C. segmentosum* and *D. pigrum* (site A specimens) are 1.14 and 0.3, respectively, while the same values for *P. melaninogenica, R. mucilaginosa* and *P. jejuni* (site B specimens) are -0.24, -0.97 and -0.06, respectively. Analysis of contrived samples containing a mixture of viral pathogen reference genomes showed classification concordance for SARS-CoV-2 (log_2_FC = -0.75) and Zika virus (log_2_FC = 0.00). Three of the viral pathogens did not pass the concordance cutoff and, therefore, warrant further investigation. The viral pathogens with discordant read counts are mammalian orthoreovirus (log_2_FC = 16.19), influenza B virus (log_2_FC = 6.83); human orthopneumovirus (log_2_FC = -3.63).

In summary, we have detected multiple microbial species, including viral pathogens and commensal bacteria, from the clinical nasal swabs analyzed in this study, with classification supported by two orthogonal methods. By generating contrived samples with mock communities of viral pathogens, we were able to detect additional viral pathogens of interest.

### CRISPR-NGS diagnostic assay performance and pathogen-host reporting

We performed an assessment of the CRISPR-NGS approach by comparing diagnostic results of SARS-CoV-2 against RT-qPCR-derived C_t_ data and clade assignments for the virus. We also aimed to determine the potential of pathogen-host genetic information that can be extracted for reporting and downstream analysis, such as data regarding co-infections, the nasal microbiome profile, antimicrobial resistance (AMR), and host gene expression at the collection site (i.e., the nasal cavity). This was undertaken by subsampling a predetermined number of sequencing reads (i.e., 40M read pairs) from each clinical specimen and contrived sample, re-mapping reads to the viral and bacterial genomes of interest and computing genome coverage breadth and depth metrics. For SARS-CoV-2, the analysis also included constructing a viral consensus genome for each sample for clade analysis.

Detectable SARS-CoV-3 read counts (out of 40 M read pairs) were averaged across samples in three ranges of C_t_ values and on average a 6-fold increase in read counts was observed, 6.4-fold for C_t_ < 23, 7.1-fold of C_t_ 23 – 30, and 4.7-fold of C_t_ 30 - 39 (Figure 4). For SARS-CoV-2, the average genome breadth of coverage was 94% up to a C_t_ value of 30, 83% up to a C_t_ value of 35, and 61% when including all samples for site A specimens. A summary of the genome coverage metrics for SARS-CoV-2, nasal microbiome species, and viral reference pathogens in the contrived samples is shown in Supporting Table 5. Sensitivity and specificity of the CRISPR-NGS assay with respect to SARS-CoV-2 detection were measured using two genome coverage metrics (number of uniquely aligned reads and genome breadth). Thresholds for detection were determined empirically and defined as follows: genome breath coverage >=3%, and number of uniquely aligned reads >=20 per 40M read pairs sequenced. For site A specimens with C_t_ values up to 35 (93 libraries), the sensitivity and specificity of SARS-CoV-2 detection were 98.4% and 100%, respectively (Table 1). For site B specimens with C_t_ values up to 32 (24 libraries), sensitivity of SARS-CoV-2 detection was 95% and specificity was 100%. To evaluate clade assignment accuracy, we compared our consensus genome-based NextClade approach against clade identification via an independent PCR-based method (Variant-seq, PerkinElmer) performed for site A specimens. We applied a 10% genome breadth cut-off as the minimum requirement for accepting clade assignments from Nextclade. Using this criterion, the clade assignment accuracy is 100% for all samples that met these criteria. (i.e., 56 out of 56 libraries).

**Figure 4.**
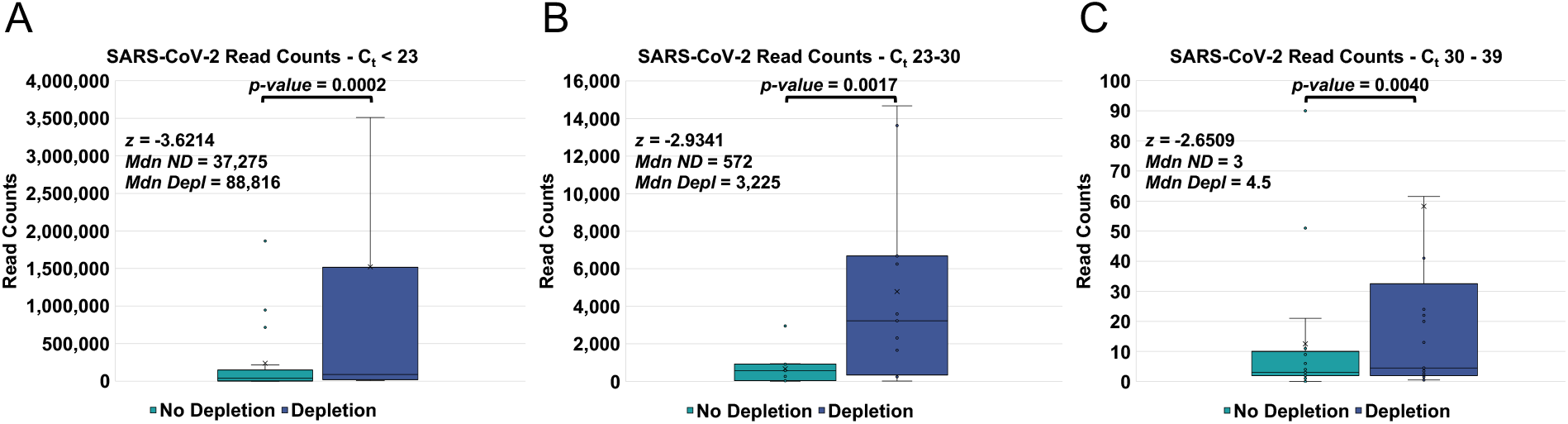
Sequencing read counts for SARS-CoV-2 in clinical specimens across Ct values. The sequencing read counts, shown on the y-axis, from Kraken2 workflow were calculated for non-depleted (blue) and depleted (purple) samples. Box and whisker plots were generated for three cycle threshold (C_t_) bins. A. C_t_ <23 (non-depleted - n = 17, depleted – n = 34). B. C_t_ 23-30 (non-depleted – n = 11, depleted – n = 22). C. C_t_ 30 – 39 (non-depleted – n = 17, depleted – n = 34). Values for the two depleted sample replicates were averaged and compared to single non-depleted samples to provide paired values for Wilcoxon Singed-Rank test. The Wilcoxon Signed-Rank Test indicated that sequence read counts to SARS-CoV-2 genome were statistically significantly higher with CRISPRclean depletion than without depletion. The z value (*z*), median of non-depleted (*Mdn ND*) and depleted (*Mdn Depl*) samples are shown in the upper left of the graph for each C_t_ bin.

We also investigated whether the proposed sequencing strategy could provide information on SARS-CoV-2 functional variants, AMR sequences, and host transcriptome responses. With respect to SARS-CoV-2, we identified the spike protein L452R mutation in one specimen. This is a mutation in the receptor-binding domain of the spike protein and is implicated in antibody resistance and immune escape (24, 25). We also identified AMR gene sequences amongst the assembled sequence contigs generated through the CZID workflow. Using AMRFinderPlus (NCBI), AMR genes were detected in a subset of the clinical specimens (n=15) from site A and B (Table 2). To identify differentially expressed (DE) host genes, we compared confirmed COVID-19 negative specimens to COVID-19 positive specimens with moderate to high viral loads (i.e., C_t_ value < 21) from site A. We identified a total of 77 up-regulated genes and 5 down-regulated genes in COVID-19 positive specimens (abs(log_2_FC) >= 1.5, and adj P < 0.05) (Supporting Table 6). Of the 82 DE genes, 19 genes overlapped with a previously identified blood-derived, interferon-stimulated gene (ISG) signature of SARS-CoV-2 infection consisting of 23 genes (26). Two interferon-inducible genes (IFI6, IFI27) from the 82 DEG list also overlapped with both the ISG signature and a blood-derived COVID-19 infection signature comprising 139 genes (26).

**Table 2.** Antimicrobial resistance (AMR) genes identified in clinical samples from Site A and B. AMR genes (Gene Symbol and Sequence Name) were identified in the assembled contigs from CZID output. The antibiotic class of each gene was determined and the number of specimens exhibiting the gene was calculated. The average percent sequence coverage and identity is shown in columns 5 and 6.

| Gene Symbol | Sequence Name | Antibiotic Class | Specimen Counts | Avg. Coverage of AMR Gene (%) | Avg. Identity AMR Gene (%) |
| --- | --- | --- | --- | --- | --- |
| aph(3')-Ia | 23S rRNA (adenine(2058)-N(6))-methyltransferase Erm(X) | Kanamycin | 6 | 70.97 | 99.43 |
| aph(3')-IIIa | aminoglycoside O-phosphotransferase APH(3')-Ia | Amikacin/<br>Kanamycin | 2 | 52.77 | 88.34 |
| cfxA3 | aminoglycoside O-phosphotransferase APH(3')-IIIa | Beta-lactam | 1 | 85.98 | 100.00 |
| erm(X) | class A extended-spectrum beta-lactamase CfxA3 | Macrolide | 1 | 60.44 | 100.00 |
| tet(37) | tetracycline efflux ABC transporter Tet(46) subunit B | Tetracycline | 1 | 55.19 | 98.75 |
| tet(W) | tetracycline resistance NADPH-dependent oxidoreductase Tet(37) | Tetracycline | 3 | 68.52 | 94.14 |
| tetB(46) | tetracycline resistance ribosomal protection protein Tet(W) | Tetracycline | 1 | 66.51 | 99.76 |
| **Total** |  |  | **15** | **67.00** | **98.41** |

## DISCUSSION

Novel technologies and strategies are sorely needed to combat infectious disease outbreaks. We believe strongly that NGS-based strategies not only have Day Zero deployment capabilities but are essential for a robust public health response at Day Zero and beyond because of the comprehensive nature of the data generated by such approaches. The issue with a NGS approach focused on RNA content is that human host derived RNA molecules, including ribosomal RNA, dominates the sequencer output and must be removed prior to sequencing. Our strategy is based on the *in vitro* application of CRISPR technology, which, because of the programmability of the CRISPR-Cas9 system, can be used to remove any known abundant and uninformative molecules from NGS libraries. The workflow involved is fast, robust, and compatible with low biomass inputs. The programmability of the CRISPR system also means that new CRISPR targets can be easily added to remove different or additional molecules and increase sensitivity. This could have considerable benefit to clinical assay development because new targets can be added with minimal alteration to the assay. In the context of infectious disease, removal of additional known human host and common bacterial sequences or contaminants should continue to improve performance up until molecular diversity is exhausted or the impact on sensitivity becomes minimal.

Although detection of the SARS-CoV-2 genome can be accomplished with legacy technology such as RT-qPCR, it is our assertion that generating many independent sequencing reads across the breadth of a pathogen genome provides more confident detection than a 100-bp amplified fragment using, for example, a fluorescent signal as output. Actual sequence information to characterize strains, clinically relevant sequence variants, co-infections, or host responses is unobtainable with focused RT-qPCR testing alone. Furthermore, although both RT-qPCR and amplicon-based targeted sequencing technologies are essential tools for detecting and tracking pathogens as they evolve, neither can meet the Day Zero requirement for a novel zoonotic pathogen.

As emphasized throughout, the mNGS strategy we have outlined in this study can be used to rapidly identify novel pathogen strains, co-infections, and host response. Several of the non-SARS-CoV-2 viruses we identified in our samples are associated with respiratory illness. One report suggests that rhinovirus will block or inhibit SARS-CoV-2 replication in lung epithelial cells by triggering an interferon response (27). This information could be critical to predict outcome or severity of disease. Mycobacterium phage was also highly prevalent in COVID-19 positive samples. Several reports suggest that severity of infection with SARS-CoV-2 could be influenced by phage therapy or stimulate trained immunity from previous mycobacterium infections and the interactions with mycobacterium phage influenced tuberculosis (28, 29). In a recently published study on co-infections and superinfections based on a literature review of papers published between October 2019 and February 2021, COVID-19 positive patients with co-infections (viral, bacterial, or fungal) had higher odds of mortality than those with COVID-19 alone (30). Many of the identified co-infections had effective treatments available.

There is still a need to validate our proposed CRISPR-NGS strategy across multiple laboratories, in multiple geographical regions, and in urban, rural, and remote settings. In this study, using samples collected and processed across multiple sites, human and bacterial RNA profiles differed between sites. These differences could be related to different methods of specimen collection, processing, and RNA extraction. To mitigate these effects, standardized methods and protocols need to be employed across labs and regions to generate comparable data. Also, if it is to be considered as deployable at scale, relevant databases harboring pathogen sequences and analysis pipelines will need to be standardized and validated. Ultimately, however, the ability of the CRISPR-NGS approach to provide a more comprehensive exploration of an individual’s infection status provides benefits that no other single approach can. We see at least six crucially important capabilities enabled as a result of this approach. First, mNGS can generate whole genome information from an RNA virus and whole transcriptome information from an active DNA viral, bacterial, or fungal infection which provides advantages over a metagenomics approach. Whole genome information enables one to track the origin and the spread of any outbreak, regardless of the sequence similarity (or lack thereof) of the target pathogen to other pathogens. Second, it can be used to detect co-infections that have the potential to increase morbidity or mortality. This includes other viral pathogens, as well as bacterial and fungal pathogens. Third, the host immune profile could be used to predict a patient’s ability to overcome infection (tolerance) or the risk of lethality from infection. Fourth, it could enable more rapid and effective identification of targets for therapeutic development. Fifth, it leverages existing and established technology with proven performance and available high throughput processing protocols. Sixth, since it has Day Zero capabilities, it will have utility immediately after the first reported case of infection and potentially mitigate the effects of the next pandemic without the need to scramble national public and private resources to meet the demands of a panicked population.

## MATERIALS AND METHODS

### Samples Used to Compare PCR and NGS Detection

For site A specimens, RNA samples were obtained from a clinical testing laboratory in California managed by PerkinElmer Scientific. Nasopharyngeal samples were extracted and tested using the PerkinElmer® SARS-CoV-2 Nucleic Acid Detection Kit. The sample set consisted of 57 COVID-positive samples with a C_t_ value for the N Gene ranging from 15.56 to 39.27 and 15 COVID-negative samples. Each sample was sequenced without CRISPR based depletion and compared to two technical replicates with CRISPR based depletion performed with approximately 13,000 single guide RNAs designed against the most abundant sequences from human and bacterial species. The final pooled library sample was quantified using the Thermo Fisher® Scientific Qubit™ HS dsDNA kit and then run on the LabChip® GX Touch™ for fragment size analysis. Samples were sequenced on an Illumina NovaSeq at 2×150bp. An 8 µL aliquot of the remaining extracted nucleic acid material from the COVID-positive samples were used as input for the NEXTFLEX® Variant-Seq™ SARS-CoV-2 v2 kit (PerkinElmer), regardless of their C_t_ value. Amplicon sequencing was completed on an Illumina® MiSeq® instrument at 2×36bp. FastQ files were uploaded to the CosmosID® SARS-CoV-2 Strain Typing Analysis Portal for analysis. SARS-CoV-2 genome coverage was also reviewed with the Integrative Genomics Viewer software (IGV). For site B specimens, nasal swabs for COVID-19 testing were collected in Arizona by TGen’s infectious disease testing facility in Flagstaff (31, 32). The remaining unused specimens were used for this study. The C_t_ values of the samples were between 14.17 and 32.02. Total RNA extraction from the nasal swab specimen was performed using the Quick-DNA/RNA MagBead kit (Zymo Research). A summary of the site A and B specimens is provided in Figure 1 and Supporting Table 1.

### Exploiting CRISPR for depletion of unwanted sequences

CRISPR-based depletion of abundant nucleic acids was performed using CRISPRclean Plus Kit (Jumpcode Genomics, Inc., San Diego) following the manufacturer’s protocols. The human rRNA CRISPR guide RNA set was designed to deplete the human mitochondrial 12S and 16S genes and human nuclear 5S, 5.8S, 18S and 28S rRNA genes, as well as the 45S precursor rRNA transcripts. The accompanying pan-bacterial rRNA CRISPR guide RNA was designed to the 5S, 16S and 23S rRNA sequences of 212 bacterial species.

### Library Construction and CRISPRclean™ depletion

For site A specimens, 10ng of each RNA sample was used as input in the PerkinElmer NEXTFLEX Rapid Directional RNA-Seq v2.0 library prep. Key steps in the library prep include first strand synthesis using random priming, second strand synthesis with uracil incorporation, fragment end-repair, adapter ligation and PCR. Prior to PCR amplification, the library is treated with Cas9 pre-complexed with guide RNA targeted to bacterial rRNA for 1 hour at 37°C followed immediately by a similar treatment with Cas9 and guide RNA targeted to human rRNA. The treatments result in the cleavage of library fragments containing rRNA sequences. A subsequent Ampure XP bead-based size selection step removes cleaved fragments and excess adapter sequences. This is followed by the standard library prep PCR to amplify the remaining (uncleaved) library. Due to material constraints, only two CRISPR-treated libraries and one mock-treated library were generated from each sample. A total of 180 libraries were produced from 60 samples. Libraries were combined into pools and loaded on 4 lanes of an Illumina NovaSeq S4 flow cell. Sequencing was performed with 2 × 150 cycles.

As for site B clinical specimens, due to limitations on RNA availability 1ng was used for library construction following the same procedure as described for site A specimens using NEXTFLEX Rapid Directional RNA-Seq Kit 2 (PerkinElmer) for cDNA library construction, and CRISPRClean kits (Jumpcode) for human and bacterial rRNA depletion. For site B contrived samples, a premixed of viral pathogen nucleic acids (ATCC virome MSA-1008) [which comprised of 4 RNA viruses; Human respiratory syncytial virus, Influenza B virus B/Florida/4/2006, Reovirus 3, and DNA viruses Human mastadenovirus F, and Human herpesvirus 5] and two SARS-CoV-2 strains (VR-1986D, VR-1992D) were purchased from ATCC. A 10-fold serial dilution was performed to span approximately 20 to 20,000 copies of the pathogens in each of the contrived sample, and in a background of 1 ng or 10 ng of human lung total RNA (Takara).

### Sequencing

For site A libraries, the concentrations were assessed through fluorometric quantification using the Qubit 4.0 (Thermo Fisher, Inc.) and the high sensitivity dsDNA assay. Library sizes were evaluated using the Agilent BioAnalyzer 2100 and the high sensitivity dsDNA kit. Libraries were normalized to 1.5 nM and were combined into 4 in the dependent pools, e.g., 45 libraries per pool, to be loaded on 4 independent lanes of an Illumina NovaSeq 6000 S4 flow cell (using a NovaSeq XP 4-Lane Kit v1.5). Sequencing was performed to produce 150 bp paired-end reads (2 × 150 bp). For site B clinical specimens and contrived samples, all libraries were sequenced on one lane of a NovaSeq 6000 S4 flow cell (2 × 150 cycles).

### Sequence Data Analysis and Interpretation Pipelines

Illumina reads generated from site A and B specimens and samples were analyzed using a unified workflow described as follows.

#### Microbiome taxonomy abundance

The 150bp paired reads were demultiplexed according to sample barcodes. The Illumina sequencing adapters were removed, and low-quality bases were trimmed using AdapterRemoval (v2.3.1). After the trimming, any reads shorter than 75bp were discarded along with their mate reads. Prior to running Kraken2-Bracken for microbial taxonomy classification, a human genome reference was built by combining GRCh38 with alternate contigs, CHM13 T2T genome (GCA_009914755.3), and the non-reference unique insertions (NUIs) identified in Wong et al (33). All trimmed reads were mapped to the human genome reference to pre-filter host reads with 95% sequence identity and 50% read length coverage as the mapping criteria. All remaining reads after host filtering were assigned taxonomy using Kraken2 (v2.1.1) with PlusPF database (release date: 1/27/2021). The domain-level and species-level taxonomic abundance values were estimated using Bracken (v2.6.0 (22)) based on the read counts from Kraken2 (21). For calculating the relative abundances among the microbial species (i.e., the microbiome space) via Kraken2-Bracken, the reads assigned to “human” were excluded from the denominator. For rRNA content estimation using Kraken2, a Kraken database (containing rRNA sequences from prokaryotes and eukaryotes) was built from the rRNA sequences collected from NCBI Nucleotide database using the following query: “biomol_rrna[PROP]” (as of March 17, 2021). For CZID-based taxonomy classification, the raw reads were uploaded to the CZID public server (pipeline v6.8), which includes its own read quality control steps. The CZID workflow performs read mapping to the NCBI non-redundant protein and nucleotide sequence databases NR and NT, respectively (the NT read mapping results were used in this analysis) and read assembly to build assembled contigs. Both sets of information were used to assign taxonomy to the input sequence read (20). The identification of microbial taxa that are likely contaminants (e.g., molecular reagents) was guided by the water blank control included in this study. A few environmental species such as Achromobacter sp. and others were considered as contaminants.

#### Genome breadth and depth coverage

Genome sequences were collected from NCBI Genbank for the top 20 species from each of the two sites based on the Kraken2-Bracken approach and the expected ATCC viral pathogen references from the contrived samples. The 40M subsampled read pairs were mapped to the combined genome sequences using BWA-MEM (v0.7.17). Only the reads with high mapping quality were kept for the downstream analysis (i.e., 95% sequence identity and 80% read length coverage). For each species, the number of mapped reads and the number of total bases mapped were collected using Bedtools (v1.9) “multicov” and Samtools (v1.9) “depth” commands, respectively, with optional parameters “-d 0 -aa” being used for Samtools “depth” command to accurately report the depths in deeply covered regions. The genome breadth coverage was calculated by the number of genome positions covered by at least one read by the total genome size. The genome depth coverage was calculated by averaging read depths across the genome.

#### SARS-CoV-2 clade identification

For the samples with 10% or higher SARS-CoV-2 genome breadth coverage, the nucleotide variants were identified, and the genome sequence was built using Bcftools (v1.9) based on the identified variants having N’s for the regions not covered by reads. The reconstructed genome sequence was used to identify SARS-CoV-2 clades using Nextclade (v1.9.0) with the downloaded dataset (tag: 2022-01-05T19:54:31Z).

#### AMR gene identification

The assembled contigs from the CZID workflow with 40M subsampled read pairs were retrieved and searched against AMR genes using NCBI AMRFinderPlus (v3.10.21).

#### Host transcriptome response

The differential gene expression and ontology enrichment analysis between COVID-19 positive (C_t_ value below 21) and confirmed negative samples from site A was done using DEGenR, an interactive Shiny app that provides integrated tools for performing differential gene expression, ranked-based ontological gene set and pathway enrichment analysis (34). Within DEGenR, the raw read counts were imported, filtered, normalized using edgeR R-package to filter out any low-expressed genes. This was followed by differential gene expression analysis using the Empirical Bayes method (eBayes) (35).

#### Functional enrichment analysis of human host response genes

To identify significant biological processes associated with the COVID-19 positive samples, we use Gene Ontology (GO) databases to assess the coherence of Differentially Expressed Genes (DEGs) (36). The Enrichr R package (37) was used to rank enriched terms among DEGs using different databases and resources, including GO biological processes. We used the Enrichr overrepresentation analysis (ORA) test incorporated (37) within DEGenR to associate biological functions to DEGs.

## Data Availability

All data produced in the present study are available upon reasonable request to the authors

## ACKNOWLEDGEMENTS

The authors would like to thank Michelle Fraser, Kerry Gunning and Amanda Cramer for providing clinical samples and Variant-Seq information for the study and Terese Hammond and Scott Layne for commenting on the approach discussed. We also want to acknowledge funding from the National Science Foundation awarded to NJS: FAIN number 2031819.

